# Challenges of Deep Learning Methods for COVID-19 Detection Using Public Datasets

**DOI:** 10.1101/2020.11.07.20227504

**Authors:** Md. Kamrul Hasan, Md. Ashraful Alam, Lavsen Dahal, Md. Toufick E Elahi, Shidhartho Roy, Sifat Redwan Wahid, Robert Martí, Bishesh Khanal

## Abstract

A large number of studies in the past months have proposed deep learning-based Artificial Intelligence (AI) tools for automated detection of COVID-19 using publicly available datasets of Chest X-rays (CXRs) or CT scans for training and evaluation. Most of these studies report high accuracy when classifying COVID-19 patients from normal or other commonly occurring pneumonia cases. However, these results are often obtained on cross-validation studies without an independent test set coming from a separate dataset and have biases such as the two classes to be predicted come from two completely different datasets. In this work, we investigate potential overfitting and biases in such studies by designing different experimental setups within the available public data constraints and highlight the challenges and limitations of developing deep learning models with such datasets. We propose a deep learning architecture for COVID-19 classification that combines two very popular classification networks, ResNet and Xception, and use it to carry out the experiments to investigate challenges and limitations. The results show that the deep learning models can overestimate their performance due to biases in the experimental design and overfitting to the training dataset. We compare the proposed architecture to state-of-the-art methods utilizing an independent test set for evaluation, where some of the identified bias and overfitting issues are reduced. Although our proposed deep learning architecture gives the best performance with our best possible setup, we highlight the challenges in comparing and interpreting various deep learning algorithms’ results. While the deep learning-based methods using chest imaging data show promise in being helpful for clinical management and triage of COVID-19 patients, our experiments suggest that a larger, more comprehensive database with less bias is necessary for developing tools applicable in real clinical settings.

## 1 Introduction

Coronavirus disease (COVID-19) that progresses and transmits rapidly has become a global pandemic with 43 million confirmed cases and 1.1 million deaths as of October 28, 2020 [1]. Early and easy detection of the disease is instrumental in saving lives and controlling the pandemic. The current gold standard for COVID-19 diagnosis is the Reverse Transcription Polymerase Chain Reaction (RT-PCR) test using respiratory specimens. However, the RT-PCR test can be affected by low viral load, has a longer turnaround time, and is still not widely available in resource-constrained areas of low-income countries [2]. The antigen tests are faster but have low sensitivity [3].

Recently, several studies have been published showing the common characteristics for COVID-19 cases in radiographic images (chest CT scans and CXRs), including bilateral, multi-focal, ground-glass opacities with a peripheral or posterior distribution, mainly in the lower lobes and early- and late-stage pulmonary consolidation [4, 5]. Thus, Artificial Intelligence (AI)-based automated tools might be useful in triage for testing and clinical management using chest radiography images [6, 7]. A large number of deep learning-based AI algorithms have been proposed in recent months for automatic classification of COVID-19 cases from normal and other pneumonia cases. These published works report high COVID-19 binary classification accuracy using either CT scans or CXRs [2, 8–19]. Although the reported metrics, such as sensitivity and specificity, are very high in most cases, these results are obtained on cross-validation studies without an independent test set coming from a separate dataset having biases, such as the two classes to be predicted from two unique datasets. AI models are likely to overfit to the distribution of training data when independent test sets are not used or are prone to learn dataset-specific artifacts rather than the actual disease characteristics.

In this work, we propose a deep learning architecture for COVID-19 classification that combines two very popular classification networks, ResNet and Xception [20, 21]. Using this proposed architecture and publicly available datasets, we design various experiments to investigate the issues of overfitting, bias, and limitations of such datasets that have been widely used for developing and evaluating COVID-19 detection algorithms in the past few months. Moreover, we compare the proposed architecture to state-of-the-art methods using an independent test set for evaluation, where some of the identified bias and overfitting issues are minimized. We validate the proposed deep learning model, trained separately for CT scans and CXRs using datasets from multiple data sources (see Table 1), and experiment its efficacy in classifying COVID-19 vs. normal, and COVID-19 vs. normal vs. other pneumonia cases. Also, we train our model using the data (PadChest [22]), which comes from a large study in Spain (for CXR), and test using the data coming from other different sources of other countries. We perform similar experiments for CT scans and demonstrate that the proposed model performs better in independent test data and generalize better than existing methods for unseen patients and test centers.

**Table 1.**
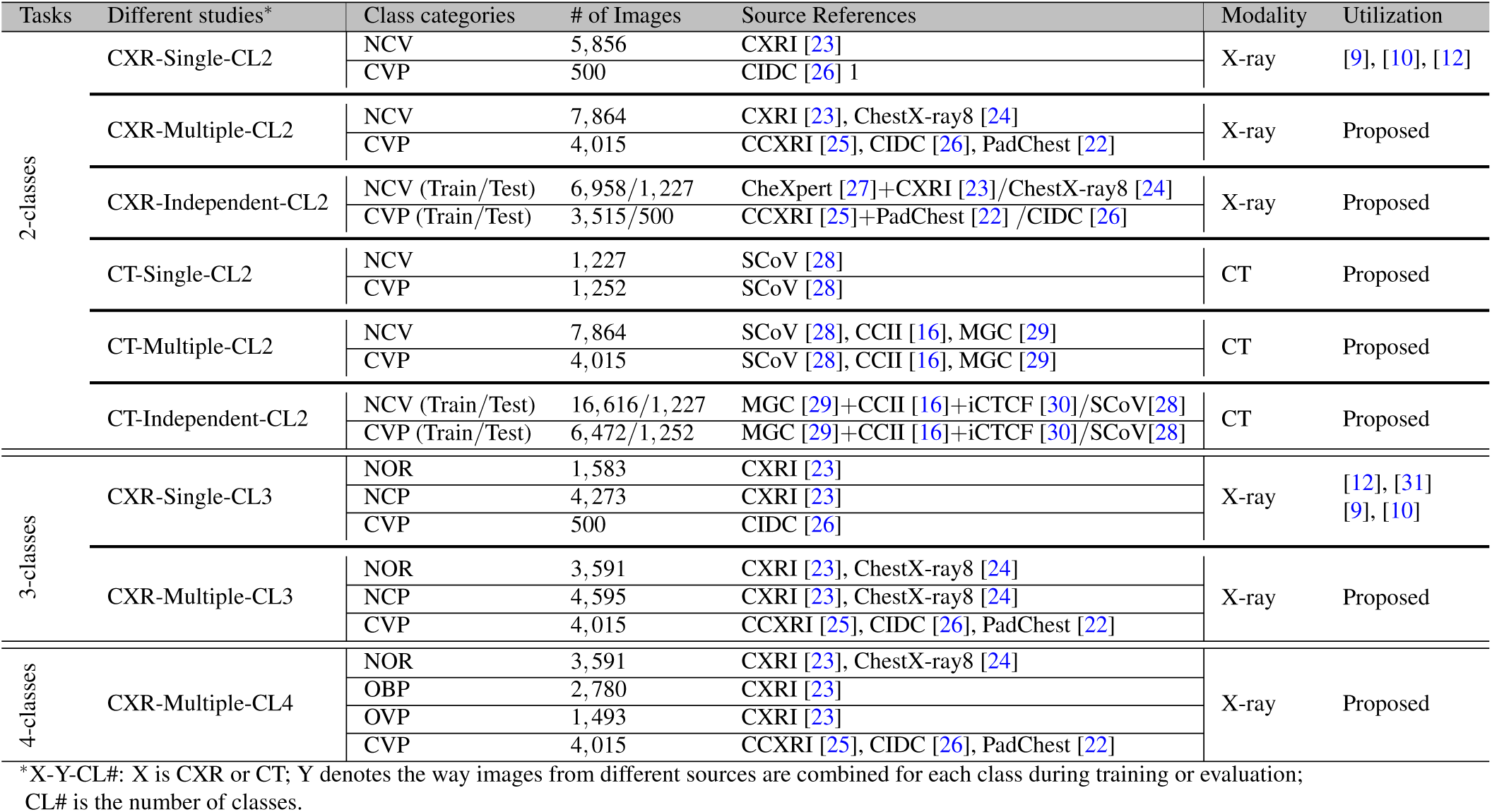
Various classification tasks utilizing CT scans or CXRs in different combinations from publicly available datasets.

In Section 2, we present the various classification tasks and study designs along with the details of different combinations of publicly available datasets for each of these designs. Section 3 presents the results followed by discussion with conclusion in Section 4. In Section 5.1, we present the proposed deep learning method in detail, followed by implementation details in Section 5.2.

### 2 Dataset and Experimental Setup

This section presents the experimental setup for various classification tasks using chest CT scans or CXRs from several publicly available datasets. The classes used for different experiments are taken from the following set:

- **NOR:** Normal; no pneumonia and COVID-19 negative
- **CVP:** COVID-19 positive pneumonia
- **OVP:** Other Viral Pneumonia; viral pneumonia but not COVID-19
- **OBP:** Other Bacterial Pneumonia; Bacteria induced non-COVID pneumonia
- **NCP:** Non-COVID Pneumonia; OBP + OVP
- **NCV:** Non-COVID; NOR + NCP

Table 1 shows the details of the experimental setup with various tasks and how various datasets are combined for these tasks. We design three different types of classification tasks: *NCV vs. CVP* (2-classes, CL2); *NOR vs. NCP vs. CVP* (3-classes, CL3); and *NOR vs. OBP vs. OVP vs. CVP* (4-classes, CL4). Several different combinations of the publicly available datasets are used for chest CT scans (labeled CT) and for chest X-rays (labeled CXR) [16, 22–30]. For each binary (CL2) or multi-class (CL3/CL4) classification tasks, we design experiments to study the impact of having single separate vs. multiple mixed sources of data for individual classes during training, labeled *Single* and *Multiple*, respectively. The setup where the test set contains images from an independent source whose images are never used during training and validation is labeled as *Independent*. Some of these setups were used in other recent studies for COVID-19 recognition, such as CXR-Single-CL2 in [9, 10, 12] and CXR-Single-CL3 in [9, 10, 12, 31]. It is worth noting that due to a lack of publicly available images, some of the designs were not possible, for example, CT-Multiple-CL3 and CT-Multiple-CL4.

In *CXR-Single-CL2*, the negative COVID CXR images (NCV) come from CXRI [23], while the positive COVID-class (CVP) comes from a separate dataset, CIDC [26]. To add more variation in each class, in *CXR-Multiple-CL2*, we add more images from other datasets resulting in 7864-NCV CXR images coming from ChestX-ray8 [24] and 4015-CVP CXR images from CCXRI [25] and PadChest [22]. To evaluate the network’s performance in distinguishing various pneumonia types, we design *CXR-Multiple-CL3* and *CXR-Multiple-CL4* where we have the same total number of images as in CXR-Multiple-CL2, but the NCV class is further split into individual pneumonia types. Similar to CXR, we use publicly available CT scan datasets. Most of these datasets contained manually selected 2D slices instead of full 3D volumes. Hence, all of the CT images referred to in this paper is 2D slices of CT scans. In *CT-Single-CL2*, we utilize 1227-NCV and 1252-CVP samples of CT scan from SCoV [28], while we have multiple sources to each class in *CT-Multiple-CL2* with 7864-NCV and 4015-CVP samples of CT scan coming from MGC [29], SCoV [28], and CCII [16].

In all of the above designs, we do not have an independent test coming from a separate dataset whose images are never used during the training. To evaluate the network’s performance on an independent test set from a separate dataset source whose images are never used during the network’s training, we design *CXR-Independent-CL2* and *CT-Independent-CL2*. For *CXR-Independent-CL2*, we have 6958-NCV images from CheXpert [27] and CXRI [23] and 3515-CVP images from CCXRI[25] and PadChest [22], while in the test set we have 1227-NCV images from ChestX-ray8 [24] and 500-CVP images from CIDC [26]. In *CT-Independent-CL2*, we build a training set with 16616-NCV and 6472-CVP samples of CT from MGC [29], CCII [16], and iCTCF [30]; and a test set with 1227-NCV and 1252-CVP samples of CT from SCoV [28].

Some examples of CT and X-ray images for different classes are presented in Fig. 1. In the setup where an independent test dataset is not available, we use 5-fold cross-validation to evaluate the performance of CVR-Net. When the independent test set is used (CXR-Independent-CL2 and CT-Independent-CL2), we evaluate the network’s performance in this independent test set. The independent setup is also used to compare the performance of the proposed method with state-of-the-art classification networks.

**Figure 1.**
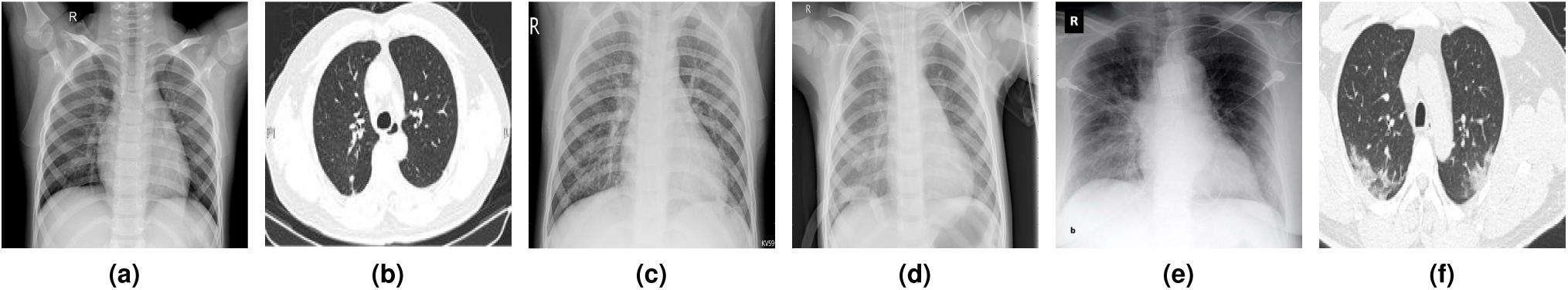
Samples of chest radiography images from the utilized datasets (a) Normal (X-ray), (b) Normal (CT), (c) Pneumonia viral (X-ray), (d) Pneumonia bacterial (X-ray), (e) COVID-19 (X-ray), and (f) COVID-19 (CT).

## 3 Results

This section presents the results of the proposed deep neural network for COVID-19 recognition, called CVR-Net (see in Section 5.1), in binary and multi-class classification tasks under various setups (see in Section 2), which highlight the impact of bias in dataset design and overfitting of the neural network. Followed by this, we compare the proposed network’s performance with state-of-the-art classification networks by training them on the same training set and using an independent test set whose images are not used during training at all.

### 3.1 Binary classification: COVID vs. Non-COVID

Table 2 presents the quantitative results of the proposed CVR-Net on the binary task: COVID-19 (CVP) vs. Non-COVID (NCV). Five-fold cross-validation results are reported with average and standard deviation, while a single value is reported when a separate test set from an independent data source is used to evaluate the results.

**Table 2.**
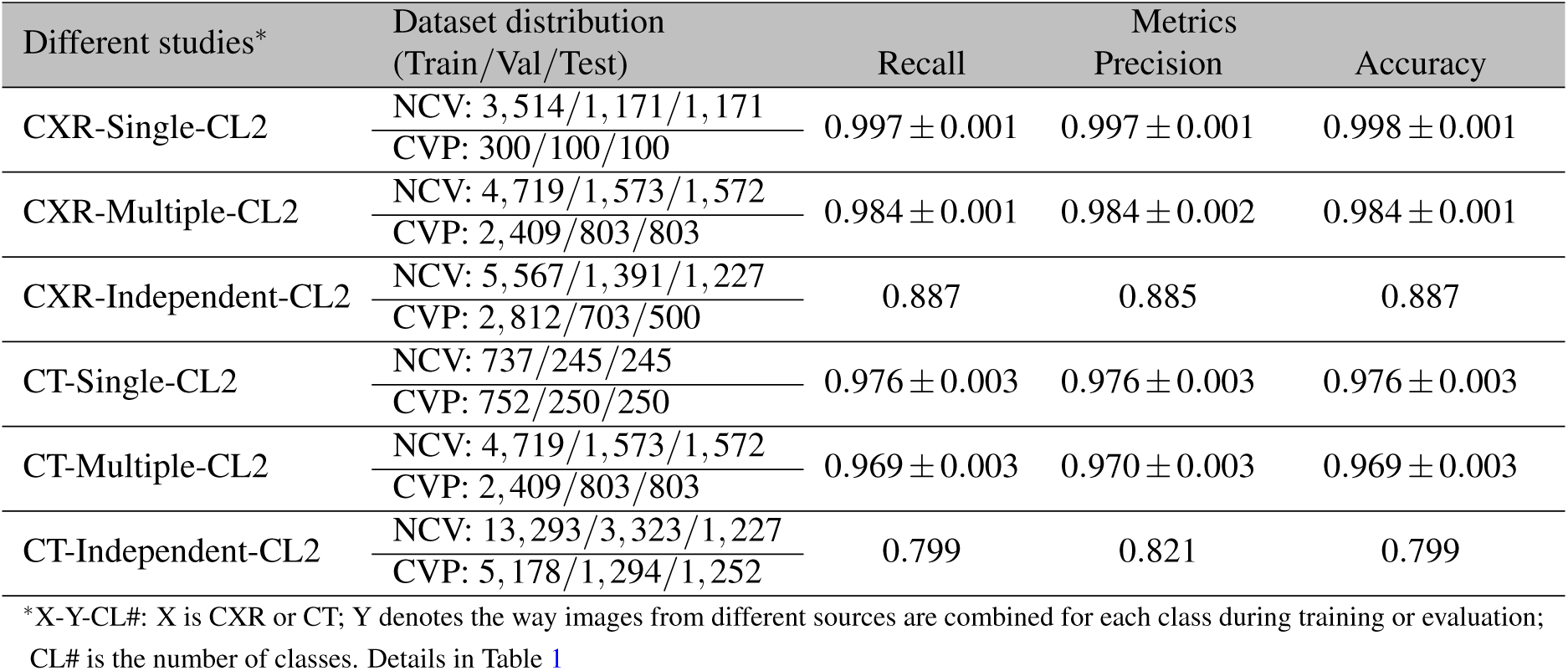
COVID-19 recognition results from different studies of binary classification (see Table 1) applying the proposed network on two different modalities of chest radiography images, wherein for single and multiple sources, we employ 5-fold cross-validation.

Table 2 demonstrates very high precision and recall in both the cases of CXR-Single-CL2 and CXR-Multiple-CL2. A slight reduction in accuracy for CXR-Multiple-CL2 compared to CXR-Single-CL2 may simply be because of relatively less overfitting to the distribution of the single particular dataset from which the individual classes were coming from in CXR-Single-CL2. As expected, the results for CXR-Independent-CL2 show reduced precision and recall, with accuracy dropping from 98 *−* 99 % in the cross-validation results to around 88 %, when using an independent test set.

The observations in the experiments with CXR is consistent in CT as well. Table 2 shows the same pattern with CT-Single-CL2 and CT-Multiple-CL2 having very high accuracy compared to CT-Independent-CL2. The cross-validation results reflect the large DL models’ overfitting nature on a relatively small dataset with limited variability of the real-world scenarios. The accuracy in CT-Independent-CL2 drops from 87 *−* 96 % in the cross-validation results to around 79 % when using the independent test set. We also notice that the accuracy with CT images is lower than CXR images.

### 3.2 Multi-class classifications: Normal, COVID, other bacterial, and viral pneumonia

Table 3 and 4 presents quantitative results of the proposed CVR-Net on two different multi-class tasks: i) 3-class problem for NOR vs. NCP vs. CVP ii) 4-class problem for NOR vs. OBP vs. OVP vs. CVP. Similar to the binary classification, cross-validation results are reported with average and standard deviation.

**Table 3.**
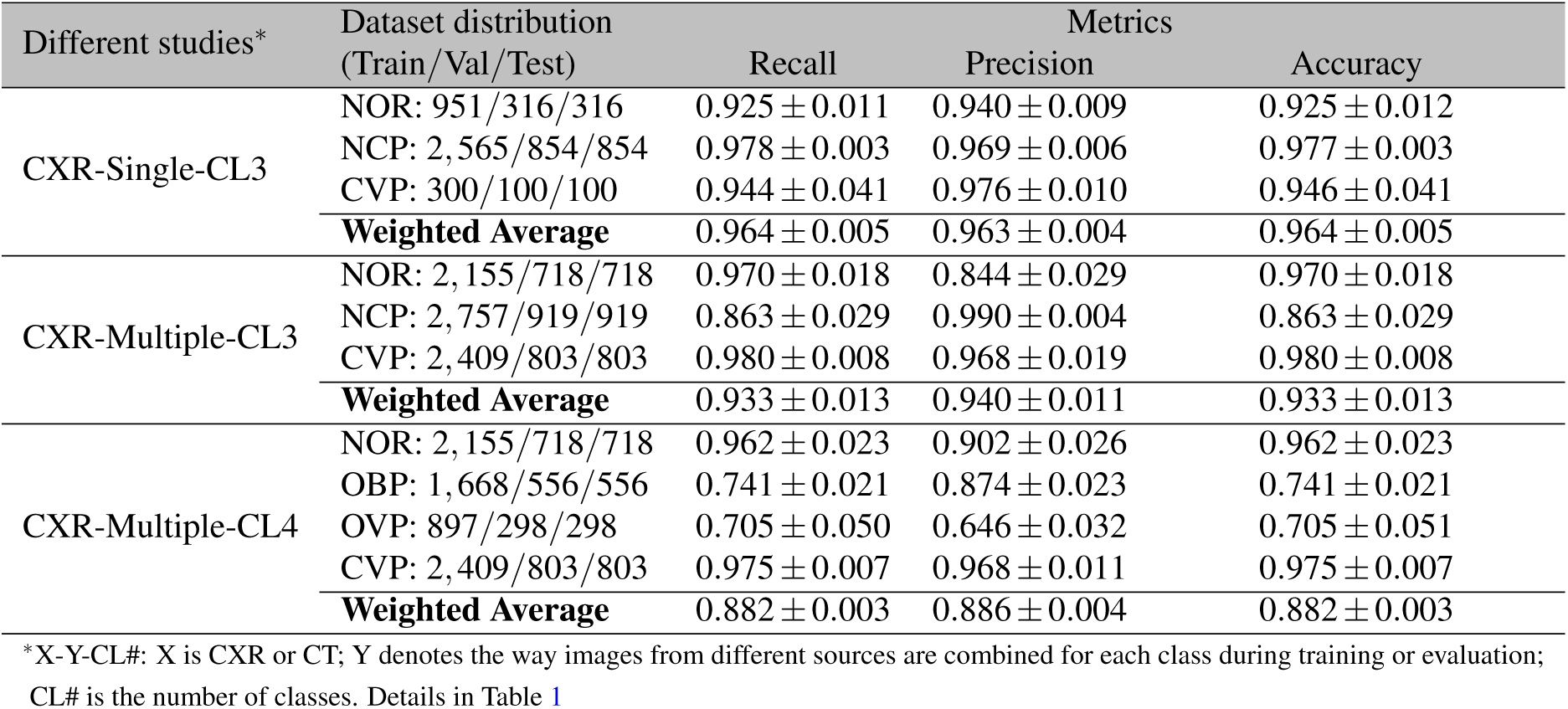
COVID-19 recognition results from different experiments of multi-class classification (see in Table 1) applying the proposed network on CXR images employing 5-fold cross-validation.

**Table 4.**
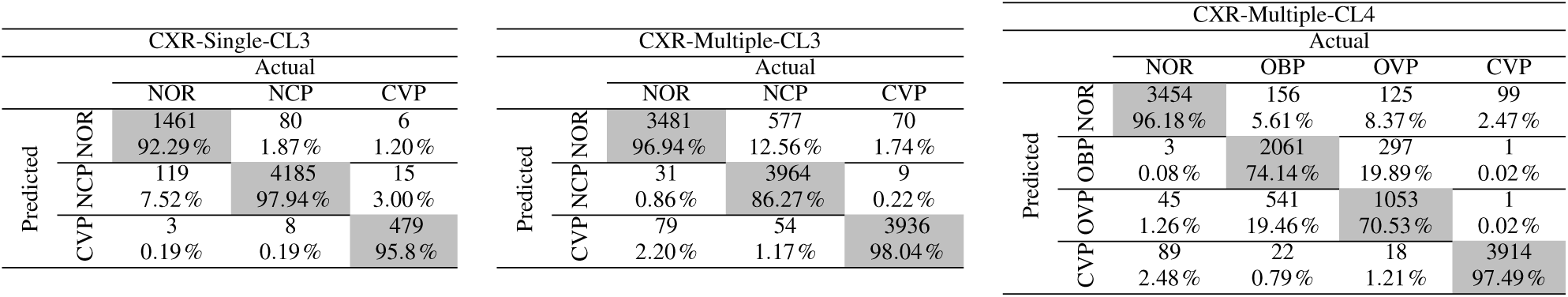
Confusion matrix for proposed networks’ results on CXR-Single-CL3, CXR-Multiple-CL3, and CXR-Multiple-CL4.

Table 4 shows that in CXR-Single-CL3, NOR and NCP rarely get predicted as CVP while a small number of CVP gets predicted as NCP and NOR. Compared to CVP, a higher fraction of NOR gets predicted as NCP. This is perhaps because the NOR and NCP classes come from the same dataset source, while CVP images are from separate sources. We see that in CXR-Multiple-CL3, fractions of NOR and CVP getting predicted as NCP are much closer. It is worth noting that NOR and NCP in CXR-Multiple-CL3 have images coming from two different datasets, but these sources still do not have the CVP images coming from separate sources. It can also be observed that adding multiple data sources in NOR and NCP has substantially increased the fraction of NCP being predicted as NOR in CXR-Multiple-CL3. From Table 2 and 3, we see that inter-fold variation is increasing with the decreased performance metrics when a new class is added with the same number of total samples when comparing CXR-Single-CL2 vs. CXR-Single-CL3 and CXR-Multiple-CL2 vs. CXR-Multiple-CL3.

In CXR-Multiple-CL4, NCP is further split into other bacterial and viral pneumonia: OBP and OVP. As seen in Table 4, the network confuses much more between OBP and OVP, both coming from the same dataset CXRI. Following the pattern of CXR-Single-CL3, we can also observe that nearly 14 % of OBP, and OVP still gets classified as NOR. CVP has relatively high precision and recall, but it is noteworthy that the source of the CVP images and the rest of the three classes do not intersect. These results further reinforce the observation in the binary classification task that seemingly high accuracy could be due to the network learning the bias in the dataset design and peculiarities of individual data sources rather than the actual underlying pathology. Unlike binary classification problems, we could not evaluate with an independent test set and perform the experiments with CT scans due to the lack of publicly available datasets for these multiple classes.

### 3.3 Comparison to the state-of-the-art

Several recent studies report the deep learning models’ performance using datasets that are not publicly available [32–34]. However, we compare these methods utilizing publicly available data using the experimental setup CXR-Independent-CL2 and CT-Independent-CL2, i.e., the setup, where test set images coming from an independent dataset whose images are never used during training of the models. Table 5 manifests the performance of the proposed CVR-Net along with other widely used and state-of-the-art classification networks and COVID-19 detection networks. The hyperparameters for all the networks used in Table 5, such as learning rate, regularizations, number of epochs, optimization algorithm, etc. are described in Subsection 5.2 at the end. The proposed CVR-Net performs the best concerning the precision, recall, and overall accuracy in both CXR and CT images. The second best is Inception-v3 for CXR and VGG-19 for CT scan.

**Table 5.** Comparison of various methods, including the proposed network (CVR-Net), where the methods are trained on the same dataset and evaluated using an independent test set, not used during training. The top three performing metrics are denoted by bold-font, underline, and double-underline, respectively.

| Methods | Parameters | CXR-Independent-CL2 |  |  | CT-Independent-CL2 |  |  |
| --- | --- | --- | --- | --- | --- | --- | --- |
|  |  | Recall | Precision | Accuracy | Recall | Precision | Accuracy |
| VGG-19 | 46M | 0.833 | 0.846 | 0.833 | <u>0.785</u> | <u>0.816</u> | <u>0.785</u> |
| Xception | 124M | <u>0.869</u> | <u>0.881</u> | <u>0.869</u> | 0.718 | 0.788 | 0.718 |
| EfficientNet-b1 | 7M | 0.832 | 0.850 | 0.832 | 0.716 | <u>0.803</u> | 0.716 |
| DenseNet-169 | 96M | 0.850 | 0.865 | 0.850 | 0.718 | 0.794 | 0.718 |
| ResNet-152 | 84M | 0.829 | 0.866 | 0.829 | 0.705 | 0.784 | 0.705 |
| Inception-v3 | 74M | <u>0.871</u> | <u>0.884</u> | <u>0.871</u> | <u>0.737</u> | 0.782 | <u>0.737</u> |
| DarkNet [9] | 1.94M | 0.712 | 0.699 | 0.712 | 0.495 | 0.245 | 0.495 |
| CoroNet [10] | 124M | <u>0.869</u> | 0.877 | <u>0.869</u> | 0.689 | 0.776 | 0.689 |
| **Proposed CVR-Net** | 48M | **0.887** | **0.885** | **0.887** | **0.799** | **0.821** | **0.799** |

Fig. 2 visualizes the regions in the input image where the neural network is activating most of its signal from when predicting COVID-19 positive class. The activation maps are shown using GradCAM with a threshold 0.6 (maximum 1) [35]. In the figure, the input images are the top three true positive images for CXR and CT having the highest sof™ax prediction output for COVID-19 class from CVR-Net. The activation map for CVR-Net as a whole is smooth and focused within the lung region, while the two branches of CVR-Net having ResNet and Xception architecture have more dispersed activation maps outside the lung region as well. This reveals that combining the two branches make the activation map more focused on the lung region. However, it is remarkably noticing that the focused region we see in the figure in the activation maps of CVR-Net does not always align with the pathology of COVID-19 seen in the CXR and CT images. For Inception, the activation maps are dispersed and smooth, but it is important to note that the images were chosen based on the highest confidence in predicting COVID-19 for CVR-Net and not for Inception.

**Figure 2.**
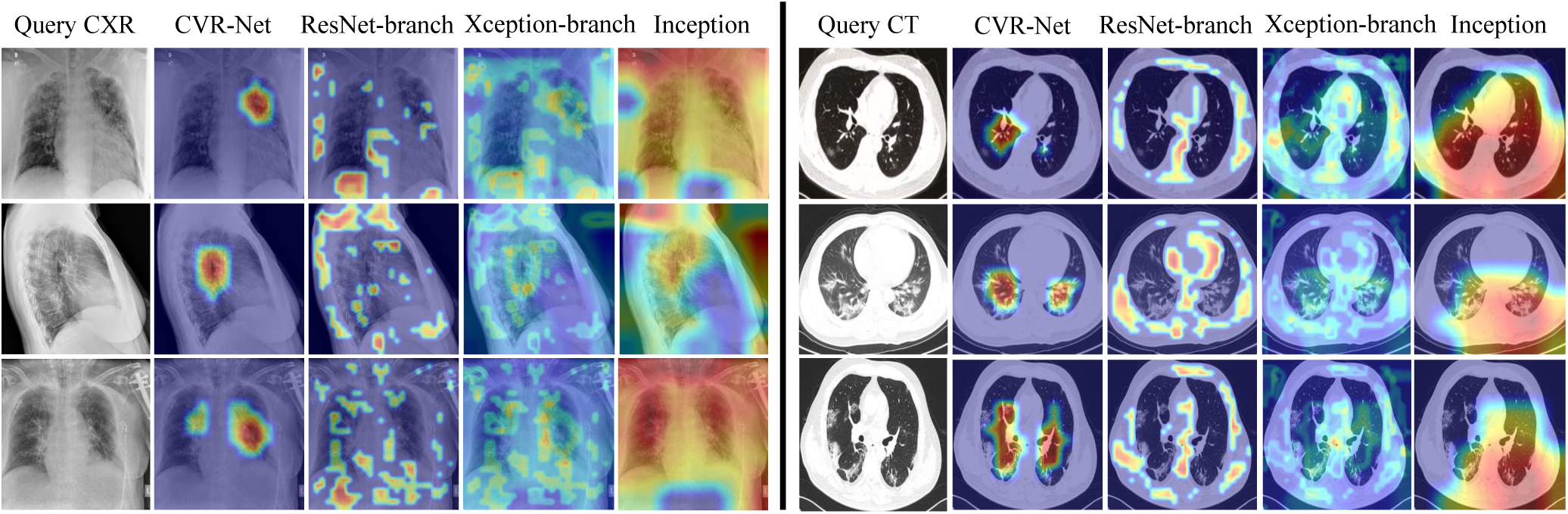
GradCAM visualizations example, showing activation map on input query CXR and CT images of COVID-19 positive class for proposed CVR-Net, encoder-1 (ResNet), encoder-2 (Xception), and Inception.

## 4 Discussion and Conclusions

In this work, we proposed a deep neural network CVR-Net for COVID-19 detection (see in Section 5.1) and employed it to conduct several experiments by utilizing publicly available CXRs and chest CT scans datasets (see in Table 1). The results show that the seemingly high accuracy reported in many recent deep learning-based algorithms for COVID-19 classification is likely due to the bias in the experimental design and overfitting. In particular, we observed two significant issues that can lead to such high accuracy that are likely not to translate to real-world settings: i) the classes to be predicted have training data coming from separate individual dataset sources. This can result in the network learning the peculiarities of the dataset from which the particular class is coming from rather than the characteristics or features of the underlying pathology. ii) the cross-validation results without an independent test set whose images are never used during training can overestimate the network’s performance. It is important to note that both the mentioned issues are common knowledge in machine learning but seems to have been overlooked or not emphasized enough in many recent works involving deep learning and COVID-19 detection [9, 10, 12, 17, 31, 36–38].

To reduce such bias and overfitting problems, we designed an experiment where the training set contains images in each class from various dataset sources, and an independent test set is used to evaluate the deep neural networks. The images from the dataset sources, which are used in the independent test set, are never used in training the model. The results show that, as expected, the performance of the deep learning model reduces in this scenario compared to the case where training and test images have images coming from the same data sources. In this more realistic setting, CVR-Net performed the best when compared against other state-of-the-art classification networks. CVR-Net (architecture detailed in section 5.1) uses multiple branches and aggregates information from different scales, creating a form of ensembling within a single network that seems to be more robust than other deep learning models such as VGG, Xception, ResNet, Inception, DenseNet, and EfficientNet, as seen in Table 5. While some of the hyperparameters such as learning rate and epochs are adapted for each model dynamically during training, we did not exhaustively optimize the hyperparameters, regularization methods, and training protocol for each of the models separately (details in section 5.2) Comparing in detail, these networks require extensive experimentation with each model to tune the hyperparameters separately and select the best regularization methods, which is outside the scope of the current paper.

Fig. 2 visualizes saliency maps for various methods. CVR-Net showing a much more concentrated map in the lung region than other networks encourages one to interpret the CVR-Net’s better capability for learning the COVID-19 feature from the chest radiography images. One approach to avoid learning from regions outside the lung area is to segment the lung and then feed only the segmented lung to the model. However, this method will then depend on the quality of the segmentation. A closer look at Fig. 2 exhibits that the heat map of CVR-Net and COVID-19 pathology in CT and chest radiograph do not always align. It is a well-known challenge in deep learning to be able to interpret such results. Although methods like GradCAM attempts to localize the most important features leading to a particular class prediction, these methods have their limitations and cannot be entirely relied upon to draw conclusive analysis [39, 40].

The experiments with 2, 3, and 4-class classification tasks show that it is quite difficult for the model to distinguish between bacterial pneumonia and other viral pneumonia. Although the results, in Table 4 for CXR-Multiple-CL4, suggest that COVID Pneumonia is well distinguished from other Pneumonia, the underlying reason is very likely that the COVID Pneumonia cases come from separate data source than the other Pneumonia. Thus, we believe that further experiments with a proper dataset are required to evaluate the model’s ability to distinguish different types of Pneumonia. We suggest that in order to evaluate the model’s ability to distinguish different classes properly, it is essential to have images for each class coming from the same settings, such as the same imaging protocol, machines, demography, *etc*. Images from multiple settings should also be included when the objective is to assess the algorithm’s ability to work on diverse settings. However, in this case, it is essential to include images from all these settings to each of the classes to reduce the bias originating from individual settings’ peculiarities.

We observed that the performance of classification results in CT-Independent-CL2 was less than CXR-Multiple-CL2. In all the experiments on CT scans, we utilized the 2D slices rather than the whole CT volume as the whole volumes are not publicly available for most experimental setups. Using the full volume instead, CT slices may be able to capture details potentially missed when the slices were manually selected. However, we cannot conclude from this study’s results that CXR is more sensitive than CT for COVID-19 diagnosis. The publicly available dataset used in the experiments come from many different sources where it is challenging to track inclusion and exclusion criteria, symptomatic vs. asymptomatic cases, and the disease severity stage at which these images were taken. Other studies have shown that CT scans are more sensitive at the early stage or in mild COVID-19 infection, but for most other symptomatic scenarios such as later stages, severe symptoms, or other comorbidities, the choice of CXR vs. CT largely depends on the specific patient conditions and the availability of the resource [41, 42].

With the limitations of the dataset used in this study, the results cannot be used to assess these networks’ performance for asymptomatic vs. symptomatic cases or various stages of the disease severity. While some studies report that CT may show single or multiple Ground Glass Opacity in the ultra-early stage of the asymptomatic cases [43], it is still unclear whether these are for only the patients who progress into the symptomatic stage, or even for those who may stay asymptomatic later on. The network application presented here may be limited to triage for prioritized RT-PCR testing from the symptomatic patients when the RT-PCR testing facility is limited or overburdened [42]. A much bigger value of these kinds of networks might be in prognostication by looking at the longitudinal imaging data [44]. For example, using longitudinal imaging data, we could develop deep learning classification models that classify symptomatic patients into: i) will have mild symptoms and can stay home ii) can develop moderate symptoms and needs to be under observation iii) can develop severe symptoms and will need ICU. We will explore this direction in our future work.

## 5 Methods

### 5.1 Proposed CVR-Net Architecture

We propose a CNN-based end-to-end network, shown in Fig. 3, that combines the convolutional and residual architecture of two popular and successful classification networks, ResNet-50 and Xception, into one with multi-scale ensembling from these two architectures [20, 21]. We remove the fully-connected layers of ResNet-50 and Xception and use the remaining layers in our two branches. We combine the information flowing through these two branches at multiple scales using concatenation and fully-connected layers. Output feature maps from two different scales of each of these two branches are fed to four Fully-connected Layer (FCL) blocks. conv3_x and conv4_x of ResNet-50, and middle-flow and entry-flow of Xception are fed to the FCL blocks that give outputs *P*_1_, *P*_2_, *P*_4_, and *P*_5_, respectively. Similarly, the output feature maps of ResNet-50 (conv5_x) and Xception (exit-flow) are first concatenated and then fed to an FCL block that gives an output *P*_3_. This allows potentially different representations learned by the two branches to be utilized by the FCL block.

**Figure 3.**
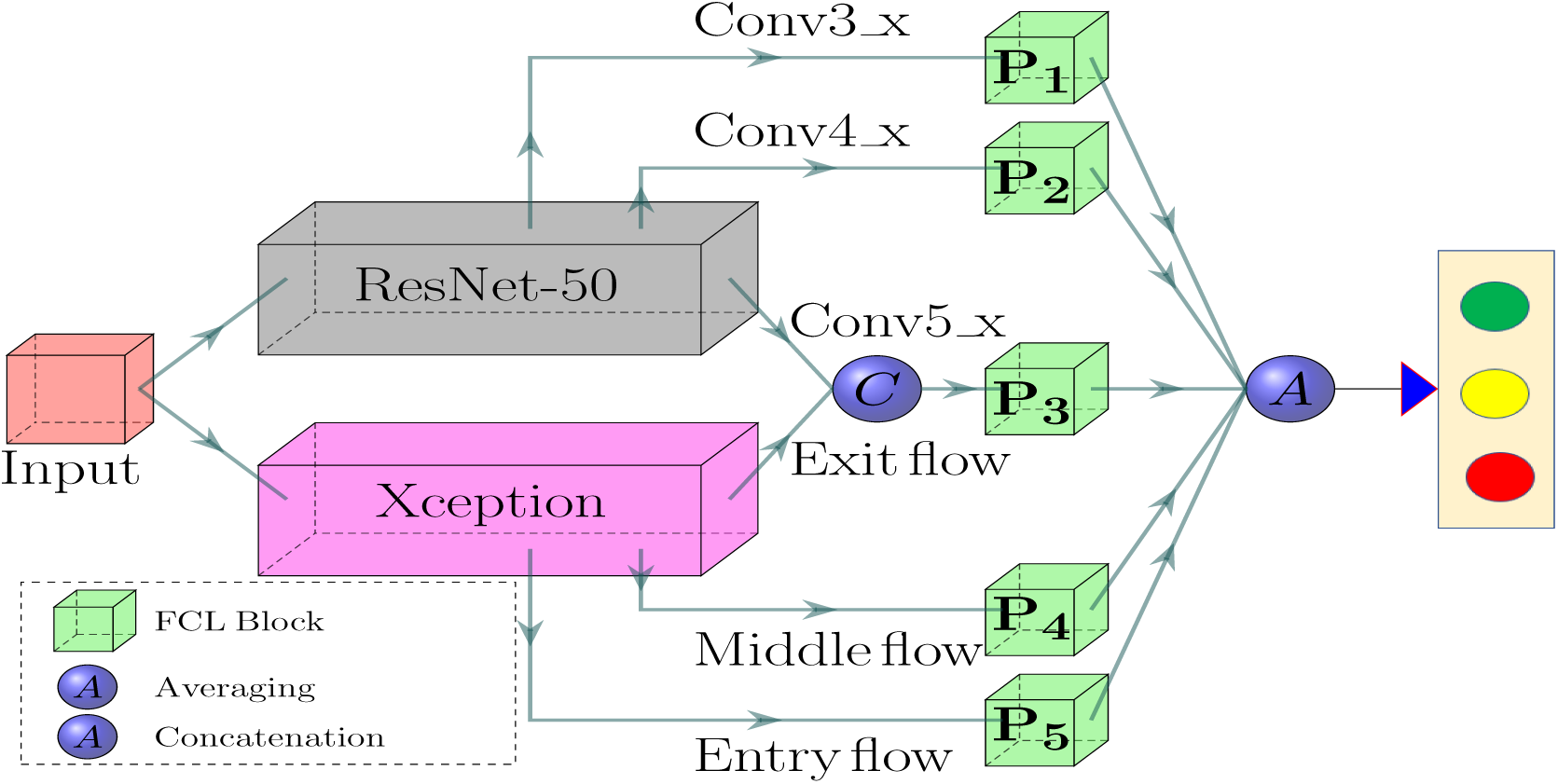
The proposed CVR-Net for the automatic COVID-19 recognition from radiography images. ResNet-50 and Xception’s convolutional layers except the fully-connected layers are used as two branches from which we combine information at multiple scales using lightweight fully-connected blocks to obtain final classification probability.

The FCL block consists of a Global Average Pooling (GAP) layer and four fully-connected layers, where the GAP layer reduces the dimension of the input feature maps [45]. In GAP, the input *h* × *w* × *d* feature map is projected to 1 × 1 × *d* vector, lending itself to a lightweight design and reducing the computational complexity for fully-connected neurons. The projected vector is then fed to a fully-connected network of 4-layers consisting of three hidden layers with 512, 128, and 64 neurons, and one output layer followed by a sof™ax layer to provide final probabilistic classification. The final prediction is the average of the sof™ax outputs from the five FCL blocks: *P*_1_ to *P*_5_. The FCL blocks are much more lightweight than the fully connected layers of ResNet-50 and Xception; thus, we have less number of parameters compared to ResNet-50 and Xception individually.

### 5.2 Training Protocol

We noticed that most of the images in all the datasets have 1 : 1 aspect ratio. Thus, we resize all the images to 224 × 224 pixels using nearest-neighbor interpolation. We apply the following stochastic augmentation on the resized images with: rotation (with probability 0.45), height & width shift (with probability 0.2), and vertical & horizontal flipping around X- and Y-axis (with probability 1), respectively. We employ categorical cross-entropy as a loss function [46], penalizing the majority class by giving higher weight in the loss function to the samples from the minority class. Each class’s weights are computed as *w*_*j*_ = *N*_*j*_*/N*, where *w*_*j*_ and *N*_*j*_ are the weight and the total number of samples for class *j* and *N* is the total number of samples.

The network weights are initialized using transfer learning [47, 48] where we use the ImageNet [49] pre-trained weights of ResNet-50 and Xception to initialize the weights of the two respective branches. We use Adam optimizer to optimize the training network with initial learning rate (*LR*), exponential decay rates (*β* _1_, *β* _2_) as *LR* = 0.0001, *β* _1_ = 0.9, and *β* _2_ = 0.999, respectively, without AMSGrad variant [50]. The initial learning rate is reduced after 12 epochs by 10.0 % if validation loss stops improving. The training is terminated after 25 epochs if the validation performance stops improving.

The models were implemented using the Python programming language and Keras framework [51] and the experiments were carried out on a machine running *Windows-10* operating system with the following hardware configuration: Intel^®^ Core™ i7-7700 HQ CPU @ 3.60 *GHz* processor with Install memory (RAM): 32.0 *GB* and GeForce GTX 1080 GPU with 8 *GB* memory.

When comparing against other state-of-the-art methods (see in Table 5), the same protocol, as described above, was used for all the networks.

## Data and Code availability

Our source codes, trained model, and sources of different utilized datasets will be made publicly available at https://github.com/kamruleee51/CVR-Net upon published.

## Data Availability

We used publicly avaliable datasets, which are free to utilize for all academic and research purposes.

## Acknowledgement

We would like to thank all the people who contributed to make the chest radiography images of COVID-19 publicly available in this current COVID-19 pandemic situation. We also thank radiologist, Dr. Ram Kumar Ghimire, for providing feedback on characteristics of COVID-19 seen in specific CXR and CT images.

## Author contributions statement

M.K.H. and B.K. conceived the experiments and study design; M.K.H wrote the primary draft of the manuscript and B.K. made the major editing; M.K.H. and M.A.A. implemented the codes and conducted the experiments; M.T.E.E, S.R., and S.R.W collected and processed the datasets; M.K.H., L.D., and B.K. analyzed the results; R.M. and B.K. supervised the research. All authors reviewed the manuscript.

## References

1. World Health Organization. WHO Coronavirus Disease (COVID-19) Dashboard (2020). https://covid19.who.int/ [Accessed: 28 October 2020].

2. Oh, Y., Park, S. & Ye, J. C. Deep learning covid-19 features on cxr using limited training data sets. IEEE Transactions on Med. Imaging (2020).

3. Scohy, A. et al. Low performance of rapid antigen detection test as frontline testing for covid-19 diagnosis. J. Clin. Virol. 104455 (2020).

4. Huang, C. et al. Clinical features of patients infected with 2019 novel coronavirus in wuhan, china. The lancet 395, 497–506 (2020).

5. Corman, V. M. et al. Detection of 2019 novel coronavirus (2019-ncov) by real-time rt-pcr. Eurosurveillance 25, 2000045 (2020).

6. Lee, E. Y., Ng, M.-Y. & Khong, P.-L. Covid-19 pneumonia: what has ct taught us? The Lancet Infect. Dis. 20, 384–385 (2020).

7. Greenspan, H., Estépar, R. S. J., Niessen, W. J., Siegel, E. & Nielsen, M. Position paper on covid-19 imaging and ai: From the clinical needs and technological challenges to initial ai solutions at the lab and national level towards a new era for ai in healthcare. Med. image analysis 66, 101800 (2020).

8. Mahmud, T., Rahman, M. A. & Fattah, S. A. Covxnet: A multi-dilation convolutional neural network for automatic covid-19 and other pneumonia detection from chest x-ray images with transferable multi-receptive feature optimization. Comput. biology medicine 122, 103869 (2020).

9. Ozturk, T. et al. Automated detection of covid-19 cases using deep neural networks with x-ray images. Comput. Biol. Medicine 103792 (2020).

10. Khan, A. I., Shah, J. L. & Bhat M. M. Coronet: A deep neural network for detection and diagnosis of covid-19 from chest x-ray images. Comput. Methods Programs Biomed. 105581 (2020).

11. Hall, L. O., Paul, R., Goldgof, D. B. & Goldgof, G. M. Finding covid-19 from chest x-rays using deep learning on a small dataset. arXiv:2004.02060 (2020).

12. Apostolopoulos, I. D. & Mpesiana, T. A. Covid-19: automatic detection from x-ray images utilizing transfer learning with convolutional neural networks. Phys. Eng. Sci. Medicine 1 (2020).

13. Apostolopoulos, I. D., Aznaouridis, S. I. & Tzani, M. A. Extracting possibly representative covid-19 biomarkers from x-ray images with deep learning approach and image data related to pulmonary diseases. J. Med. Biol. Eng. 1 (2020).

14. Rajaraman, S. et al. Iteratively pruned deep learning ensembles for covid-19 detection in chest x-rays. arXiv:2004.08379 (2020).

15. Togğaçar, M., Ergen, B. & Cömert, Z. Covid-19 detection using deep learning models to exploit social mimic optimization and structured chest x-ray images using fuzzy color and stacking approaches. Comput. Biol. Medicine 103805 (2020).

16. Zhang, K. et al. Clinically applicable ai system for accurate diagnosis, quantitative measurements, and prognosis of covid-19 pneumonia using computed tomography. Cell (2020).

17. Singh, D., Kumar, V. & Kaur, M. Classification of covid-19 patients from chest ct images using multi-objective differential evolution–based convolutional neural networks. Eur. J. Clin. Microbiol. & Infect. Dis. 1–11 (2020).

18. Huang, L. et al. Serial quantitative chest ct assessment of covid-19: Deep-learning approach. Radiol. Cardiothorac. Imaging 2, e200075 (2020).

19. Minaee, S., Kafieh, R., Sonka, M., Yazdani, S. & Soufi, G. J. Deep-covid: Predicting covid-19 from chest x-ray images using deep transfer learning. arXiv:2004.09363 (2020).

20. He, K., Zhang, X., Ren, S. & Sun, J. Deep residual learning for image recognition. In Proceedings of the IEEE conference on computer vision and pattern recognition, 770–778 (2016).

21. Chollet, F. Xception: Deep learning with depthwise separable convolutions. In Proceedings of the IEEE conference on computer vision and pattern recognition, 1251–1258 (2017).

22. Bustos, A., Pertusa, A., Salinas, J.-M. & de la Iglesia-Vayá, M. Padchest: A large chest x-ray image dataset with multi-label annotated reports. Med. image analysis 66, 101797 (2020).

23. Paul Mooney. Chest X-Ray Images (Pneumonia) (2018). https://www.kaggle.com/paultimothymooney/chest-xray-pneumonia [Accessed: 10 July 2020].

24. Wang, X. et al. Chestx-ray8: Hospital-scale chest x-ray database and benchmarks on weakly-supervised classification and localization of common thorax diseases. In Proceedings of the IEEE conference on computer vision and pattern recognition, 2097–2106 (2017).

25. Chowdhury, M. E. et al. Can ai help in screening viral and covid-19 pneumonia? arXiv:2003.13145 (2020).

26. Cohen, J. P. et al. Covid-19 image data collection: Prospective predictions are the future. arXiv:2006.11988 (2020).

27. Irvin, J. et al. Chexpert: A large chest radiograph dataset with uncertainty labels and expert comparison. In Proceedings of the AAAI Conference on Artificial Intelligence, vol. 33, 590–597 (2019).

28. Soares, E., Angelov, P., Biaso, S., Froes, M. H. & Abe, D. K. Sars-cov-2 ct-scan dataset: A large dataset of real patients ct scans for sars-cov-2 identification. medRxiv (2020).

29. Zhao, J., Zhang, Y., He, X. & Xie, P. Covid-ct-dataset: a ct scan dataset about covid-19. arXiv:2003.13865 (2020).

30. Ning, W. et al. ictcf: an integrative resource of chest computed tomography images and clinical features of patients with covid-19 pneumonia. Res. square preprint (2020).

31. Wang, L. & Wong, A. Covid-net: A tailored deep convolutional neural network design for detection of covid-19 cases from chest x-ray images. arXiv:2003.09871 (2020).

32. Harmon, S. A. et al. Artificial intelligence for the detection of covid-19 pneumonia on chest ct using multinational datasets. Nat. communications 11, 1–7 (2020).

33. Wang, Z. et al. Automatically discriminating and localizing covid-19 from community-acquired pneumonia on chest x-rays. Pattern Recognit. 107613 (2020).

34. Song, Y. et al. Deep learning enables accurate diagnosis of novel coronavirus (covid-19) with ct images. medRxiv (2020).

35. Selvaraju, R. R. et al. Grad-cam: Visual explanations from deep networks via gradient-based localization. In Proceedings of the IEEE international conference on computer vision, 618–626 (2017).

36. Sethy, P. K. & Behera, S. K. Detection of coronavirus disease (covid-19) based on deep features. Preprints 2020030300, 2020 (2020).

37. Hemdan, E. E.-D., Shouman, M. A. & Karar, M. E. Covidx-net: A framework of deep learning classifiers to diagnose covid-19 in x-ray images. arXiv:2003.11055 (2020).

38. Narin, A., Kaya, C. & Pamuk, Z. Automatic detection of coronavirus disease (covid-19) using x-ray images and deep convolutional neural networks. arXiv:2003.10849 (2020).

39. Adebayo, J. et al. Sanity checks for saliency maps. In Advances in Neural Information Processing Systems, 9505–9515 (2018).

40. Reyes, M. et al. On the interpretability of artificial intelligence in radiology: Challenges and opportunities. Radiol. Artif. Intell. 2, e190043 (2020).

41. Rubin, G. D. et al. The role of chest imaging in patient management during the covid-19 pandemic: a multinational consensus statement from the fleischner society. Chest (2020).

42. Cleverley, J., Piper, J. & Jones, M. M. The role of chest radiography in confirming covid-19 pneumonia. bmj 370 (2020).

43. Gozansky, E. K. & Moore, W. H. Sars-cov-2 from the trenches: a perspective from new york city. Am. J. Roentgenol. 1–2 (2020).

44. Rousan, L. A., Elobeid, E., Karrar, M. & Khader, Y. Chest x-ray findings and temporal lung changes in patients with covid-19 pneumonia. BMC Pulm. Medicine 20, 1–9 (2020).

45. Lin, M., Chen, Q. & Yan, S. Network in network. arXiv:1312.4400 (2013).

46. Zhang, Z. & Sabuncu, M. Generalized cross entropy loss for training deep neural networks with noisy labels. In Advances in neural information processing systems, 8778–8788 (2018).

47. Shin, H.-C. et al. Deep convolutional neural networks for computer-aided detection: Cnn architectures, dataset characteristics and transfer learning. IEEE transactions on medical imaging 35, 1285–1298 (2016).

48. Tajbakhsh, N. et al. Convolutional neural networks for medical image analysis: Full training or fine tuning? IEEE transactions on medical imaging 35, 1299–1312 (2016).

49. Krizhevsky, A., Sutskever, I. & Hinton, G. E. Imagenet classification with deep convolutional neural networks. In Advances in neural information processing systems, 1097–1105 (2012).

50. Kingma, D. P. & Ba, J. Adam: A method for stochastic optimization. arXiv:1412.6980 (2014).

51. Chollet, F. Keras. https://github.com/fchollet/keras (2015).

